# Applying Machine Learning to Blood Count Data Predicts Sepsis with ICU Admission

**DOI:** 10.1101/2022.10.21.22281348

**Authors:** Paul C. Ahrens, Daniel Steinbach, Maria Schmidt, Martin Federbusch, Lara Heuft, Christoph Lübbert, Matthias Nauck, Matthias Gründling, Berend Isermann, Sebastian Gibb, Thorsten Kaiser

## Abstract

**Background:** Delay in diagnosing sepsis results in potentially preventable deaths. Mainly due to their complexity or limited applicability, machine learning (ML) models to predict sepsis have not yet become part of clinical routines. For this reason, we created a ML model that only requires complete blood count (CBC) diagnostics.

**Methods:** Non-intensive care unit (non-ICU) data from a German tertiary care centre were collected from January 2014 to December 2021. Patient age, sex, and CBC parameters (haemoglobin, platelets, mean corpuscular volume, white and red blood cells) were utilised to train a boosted random forest, which predicts sepsis with ICU admission. Two external validations were conducted using data from another German tertiary care centre and the Medical Information Mart for Intensive Care IV database (MIMIC-IV). Using the subset of laboratory orders also including procalcitonin (PCT), an analogous model was trained with PCT as an additional feature.

**Findings:** After exclusion, 1,381,358 laboratory requests (2016 from sepsis cases) were available. The derived CBC model shows an area under the receiver operating characteristic (AUROC) of 0.872 (95% CI, 0.857–0.887) for predicting sepsis. External validations show AUROCs of 0.805 (95% CI, 0.787–0.824) and 0.845 (95% CI, 0.837–0.852) for MIMIC-IV. The model including PCT revealed a significantly higher performance (AUROC: 0.857; 95% CI, 0.836–0.877) than PCT alone (AUROC: 0.790; 95% CI, 0.759–0.821; p<0.001).

**Interpretation:** Our results demonstrate that routine CBC results could significantly improve diagnosis of sepsis when combined with ML. The CBC model can facilitate early sepsis prediction in non-ICU patients with high robustness in external validations. Its implementation in clinical decision support systems has strong potential to provide an essential time advantage and increase patient safety.

**Funding:** The study was part of the AMPEL project (www.ampel.care), which is co-financed through public funds according to the budget decided by the Saxon State Parliament under the RL eHealthSax 2017/18 grant number 100331796.

## Introduction

Sepsis is the cause of an estimated 20% of deaths worldwide.^1^ Early detection and treatment of sepsis substantially improves outcome.^2–4^ Several screening tools have been developed to identify septic patients in a timely fashion. Score-based early warning systems, such as Systemic Inflammatory Response Syndrome (SIRS), quick Sepsis-related Organ Failure Assessment (qSOFA), and National Early Warning Score (NEWS), lack either specificity or sensitivity outside the intensive care unit (ICU) and are usually performed only in cases of clinical suspicion.^4–6^ Though numerous biomarkers (over 250 by 2019) associated with sepsis have been identified, most are poorly evaluated for their role in clinical practice.^7^ Only procalcitonin (PCT) is implemented in current clinical sepsis guidelines, but its use is recommended only for discontinuation of antibiotic therapy.^4^

Screening tools based on machine learning (ML) have outperformed the traditional screening methods mentioned above in detecting and predicting sepsis, when focused on ICU data.^8,9^ Nearly all these models require various input features, including vital signs and clinical observations, that are often not available in electronic health records (EHRs), particularly in low- and middle-income countries where the incidence and mortality of sepsis is also high.^1,9,10^ A suitable prediction tool should predict sepsis early, ideally before ICU admission, and require only easily accessible clinical data. However, no corresponding ML model trained only on non-ICU data and not relying on vital signs has yet been published. With high applicability even in situations with missing further clinical information in mind, we describe here the application of ML to develop a precise prediction model from complete blood counts (CBCs) that enables early and reliable prediction of patients at risk of sepsis and ICU admission.

External validation is crucial for the assessment of model performance.^11^ In our study, we validated the model on an internal hold-out dataset, a large independent cohort from another tertiary care centre and the Medical Information Mart for Intensive Care IV database (MIMIC-IV). This dataset has been recently published and is the only public dataset that includes non-ICU data.^12^ To enable future development of related models, we make our data publicly available, which contains over 850,000 admissions to non-ICU wards.

## Methods

In this retrospective cohort study, we included all in- and outpatients aged 18 or older of the University of Leipzig Medical Centre (UML) between January 2014 and December 2021, for whom we acquired age, sex, ICD-10 coded diagnoses, and, if applicable, time of admission to an ICU. Additionally, we collected the results of CBCs taken on all non-ICU wards and outpatient clinics. Blood samples were taken on normal wards on behalf of the attending physicians either routinely (in general, two to three times a week) or based on acute orders. Patients with incomplete CBCs or without a CBC were excluded.

The Ethics Committee at the Leipzig University Faculty of Medicine approved the study (reference number: 214/18-ek). This study is published in accordance with the Transparent Reporting of a multivariable prediction model for Individual Prognosis or Diagnosis (TRIPOD) statement.^13^

### Data Processing and Model Development

All data preprocessing was performed using R version 4.2.0 and the data.table R package version 1.14.2.^14,15^ Summary tables were built using the gtsummary R package version 1.6.0.^16^ The ML was carried out using MATLAB version 9.7.0.1247435 (R2019b).^17^

First, we divided the study data into control and sepsis groups according to sepsis-related ICD-10 codes. In the sepsis group, only CBCs from patients with ICU stay and admission were labelled. In sepsis cases with multiple admissions to ICUs, we expected the first ICU episode to be sepsis-related and excluded all subsequent CBCs. To avoid systemic bias of our results due to postoperative sepsis development and SIRS caused by the preceding interventions, we included only admissions to medical ICUs and excluded all cases that were explicitly coded as SIRS. As incomplete CBCs are rare in daily practice, we chose complete-case analysis. The resulting dataset was then split into a training dataset (UMLT 2014-2019) and a hold-out validation set (UMLV 2020-2021). All inclusion criteria are depicted in Figure 1.

**Figure 1:**
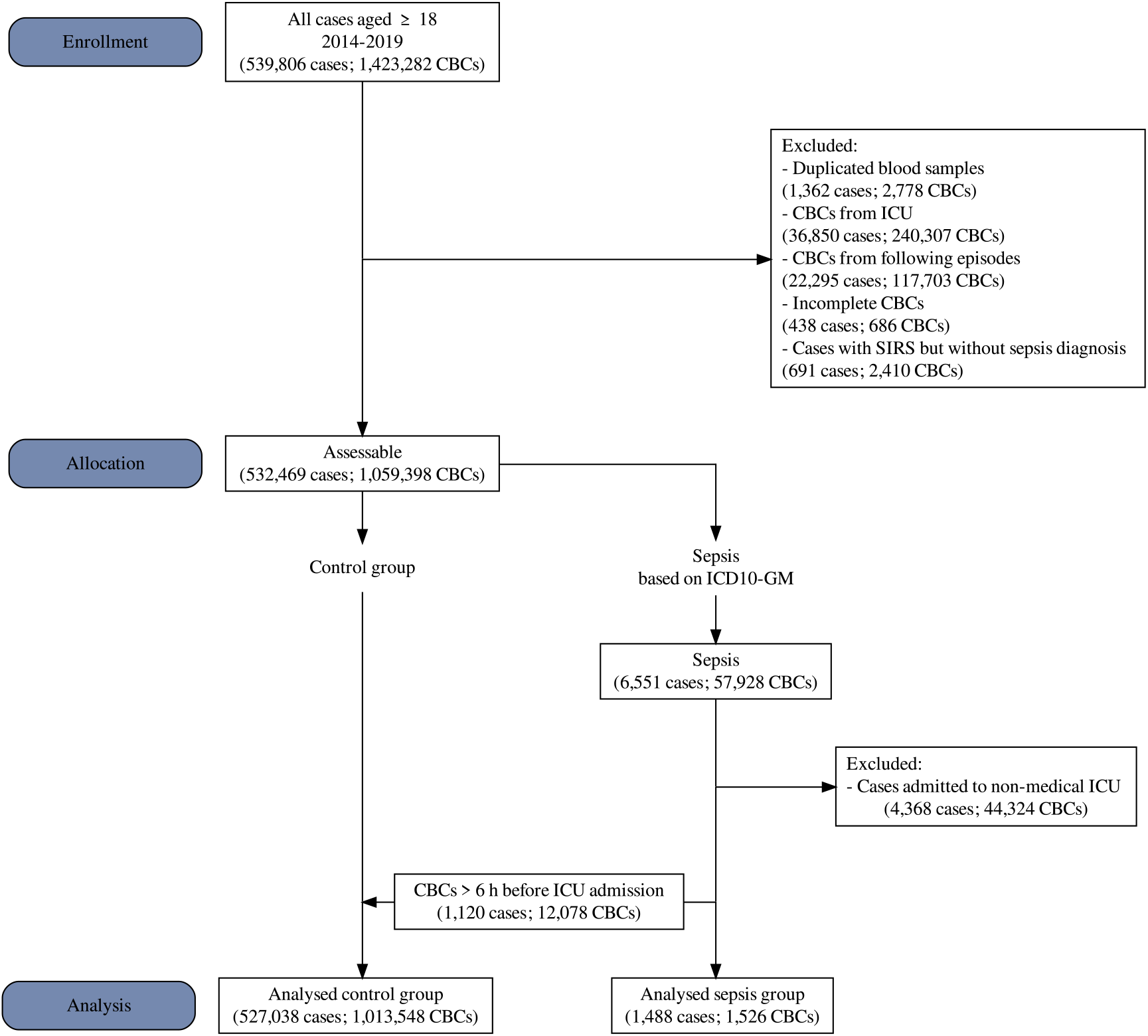
The flowchart depicts the inclusion criteria and the size of the main dataset from UMLT. UMLT: University of Leipzig Medical Center (Training Set); CBC: complete blood count; ICU: intensive care unit; SIRS: systemic inflammatory response syndrome; ICD-10-GM: International Statistical Classification of Diseases and Related Health Problems Version 10 – German Modification

We used the control or sepsis label as the response variable and age, sex, haemoglobin (HGB), mean corpuscular volume (MCV), platelets (PLTs), red blood cells (RBCs), and white blood count (WBC) as feature variables to train a random forest model. This selection of parameters was based on a laboratory medicine rationale, as these parameters represent the main characteristics of the CBC. For training, we used the RUSBoost algorithm,^18^ which is a hybrid sampling and boosting algorithm for ensemble learning. The random under sampling (RUS) balances the control and sepsis cases for the training of every tree by randomly selecting a sample of control cases equal to the number of sepsis cases. While still selecting one control CBC per sepsis CBC we oversampled the pre-septic CBCs that were taken more than 6 hours before ICU admission in the control group by a factor of 10. For hyperparameter optimisation a 5-fold cross-validation was performed. All CBCs were evaluated independently of previous or subsequent measurements. The prediction performance of our CBC model on all datasets was assessed using the area under the receiver operating characteristic (AUROC). To assess prediction performance as a function of time before ICU admission, separate AUROCs were calculated for different time intervals. In each of these intervals, CBCs of sepsis cases were labelled and used as response variable whereas subsequent CBCs were excluded. Clinically relevant sensitivity cut-offs of 90% and 50% were used for the calculation and illustration of specificities.

### Model Validation

To validate the CBC model, we tested it on the hold-out validation set (UMLV 2020-2021) and two external datasets.

First, we acquired an equivalent dataset of all patients aged 18 or older admitted to the University Medicine Greifswald (UMG) between January 2015 and December 2020. We applied the same exclusion and data preprocessing steps as described above for the UML data. The Ethics Committee at UMG approved the study (reference number: BB133/10).

Second, we utilised the recently published and publicly available MIMIC-IV dataset.^12^ Before applying the same exclusion and data preprocessing steps as described above, we labelled the patients with sepsis based on ICD-9 and -10 codes, since only some of the cases in the MIMIC-IV dataset were coded using ICD-10.

### PCT Comparison

To evaluate the performance of a trained prediction model in comparison with the biomarker PCT, we selected from the UML dataset all laboratory orders in which PCT was also measured. On the UMLT subset we trained a new CBC+PCT model analogous to the CBC model with PCT as an additional biomarker. To avoid overfitting the model due to the significantly smaller dataset, we limited the number of splits to 10 per tree. Statistical significance for the comparison was determined using the DeLong test.^19^ An alpha level of 0.05 was considered statistically significant.

More detailed information on the methods used in this study is presented in the supplement.

## Results

### Training and Internal Validation

At UML, 1,934,343 CBC measurements were performed in 724,059 cases from January 2014 to December 2021. The final UMLT dataset provided 1,013,548 CBCs (527,038 cases) in the control group, and 1,526 CBCs (1,488 cases) in the sepsis group, which met the inclusion criteria to train the ML model. More detailed information is displayed in Figure 1.

The sepsis sample consisted of 938 (63%) male and 550 (37%) female cases, with a median age of 67 (IQR, 57–76). The sepsis and control group differed significantly in all characteristics (see supplement for tabular comparisons). In 686 (0.05%) CBCs, at least one parameter was missing.

On the UMLV hold-out dataset, the AUROC of the derived CBC model was 0.872 (95% CI, 0.857–0.887, Figure 2). Adjusting the model to a sensitivity of 90% (S90), specificity was 65.1% (95% CI, 64.9–95.2); with a sensitivity of 50% (S50), specificity reached 95.2% (95% CI, 95.1–95.3). Corresponding to the sepsis label prevalence of 0.13% present in the final dataset, the positive predictive value (PPV) for S50 was 1.37% (95% CI, 1.2–1.55), and the negative predictive value (NPV) was 99.93% (95% CI, 99.92–99.94), versus PPV 0.34% (95% CI, 0.31– 0.38) and NPV 99.98% (95% CI, 99.97–99.98) for S90. Cohort analysis revealed the highest prevalence (0.81%) and PPV (5.2%; 95% CI, 4.46–6.03; S50) in the emergency department. Additional results of the cohort analysis and an illustration of the dependence of PPV on the prevalence and sensitivity can be found in the supplement.

**Figure 2:**
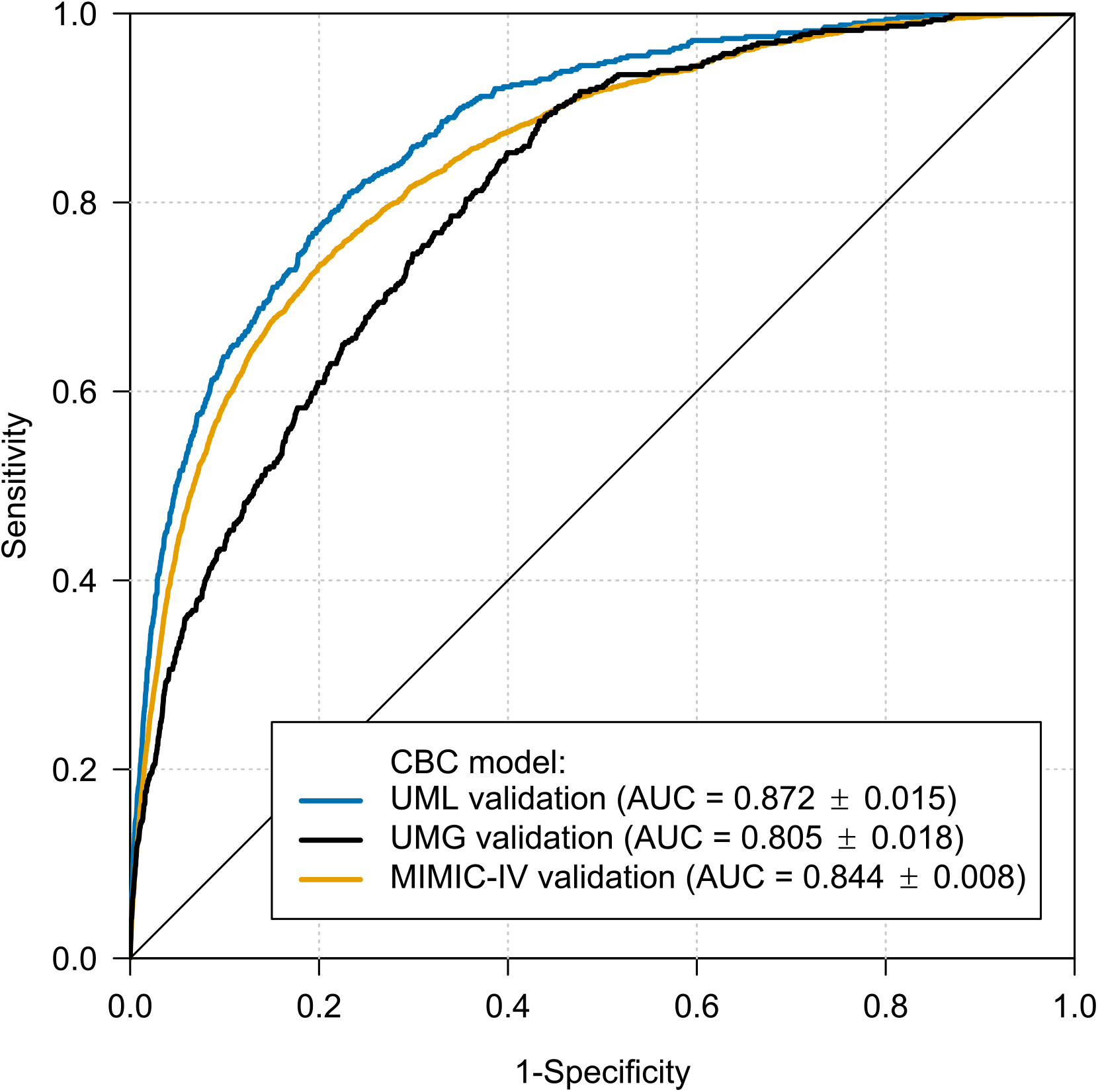
ROC curves of the CBC model for internal and external validation data. ROC:receiver operating characteristic; AUC: area under the receiver operating characteristic with 95% confidence intervals; CBC: complete blood count; UML: University Medicine Leipzig; UMG: University Medicine Greifswald; MIMIC-IV: Medical Information Mart for Intensive Care IV database.

The AUROCs decreased with time interval to ICU admission to 0.848 for CBCs taken 6-12 h before ICU admission, 0.820 for 12-24 h, 0.799 for 24-48 h, 0.774 for 2-7 d and to a baseline AUROC of 0.721 for CBCs ordered 28 days or more before ICU admission (Figure 3). In addition to these temporal dynamics, significantly different score values were calculated between patients with and without sepsis diagnosis admitted to an ICU (Figure S2, supplement). Except for sex, all parameters showed relevant importance in the resulting ML model. WBC was found to have the largest impact on the prediction (Figure S8, supplement).

**Figure 3:**
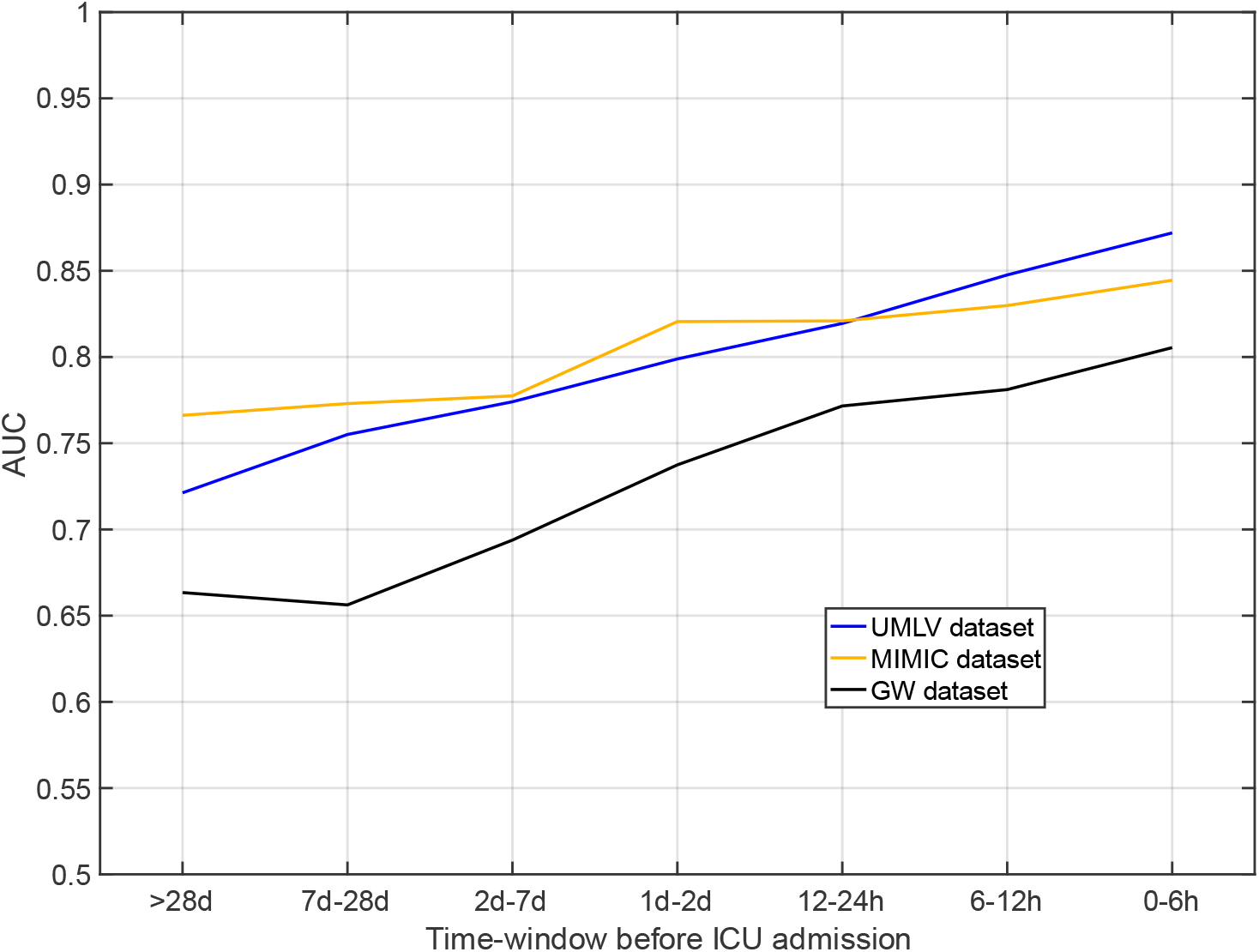
Temporal dynamics of the CBC model. AUC: area under the receiver operating characteristic; CBC: complete blood count; ICU: intensive care unit, UMLV: University of Leipzig Medical Center (Validation Set); UMG: University Medicine Greifswald; MIMIC-IV: Medical Information Mart for Intensive Care IV database.

### External Validations

The CBC model was tested on data from two large external datasets (UMG and MIMIC-IV). The UMG dataset consisted of 645,874 CBC measurements from 169,058 cases, performed between January 2015 and December 2020. With analogous application of the UML exclusion criteria, the final validation dataset included 437,629 CBCs (157,680 cases) in the control group and 448 CBCs (438 cases) in the sepsis cohort. The AUROC of the ML algorithm was 0.805 (95% CI, 0.787–0.824).

MIMIC-IV provides data from over 40,000 patients who stayed in ICUs of the Beth Israel Deaconess Medical Center (Boston, USA) from 2008 to 2019. In total, it contains 3,322,100 CBCs from 586,743 cases. After the exclusion process, 2,511,592 CBCs (559,135 cases), including 2,638 with positive sepsis labelling (2,513 cases), were available for validation. The AUROC of the CBC model (trained only with the UMLT data) remained high at 0.845 (95% CI, 0.837–0.852). Clinical and sociodemographic characteristics of the sepsis cohorts in the three datasets are displayed in Table 1.

**Table 1:** Baseline characteristics of patients with sepsis in UMLT, UMLV, UMG, and MIMIC-IV.

| Characteristic | Overall<sup>1</sup> | UMLT<sup>1</sup> | UMLV<sup>1</sup> | UMG<sup>1</sup> | MIMIC-IV<sup>1</sup> | p-value<sup>2</sup> |
| --- | --- | --- | --- | --- | --- | --- |
| General |  |  |  |  |  |  |
| N (cases) | 4,911 (100.0) | 1,488 (30.3) | 472 (9.6) | 438 (8.9) | 2,513 (51.2) |  |
| Age | 68 (58 – 79) | 67 (57 – 76) | 68 (59 – 78) | 69 (58 – 78) | 69 (58 – 81) | <0.001 |
| Male | 2,961 (60.3) | 938 (63.0) | 310 (65.7) | 278 (63.5) | 1,435 (57.1) | <0.001 |
| Female | 1,950 (39.7) | 550 (37.0) | 162 (34.3) | 160 (36.5) | 1,078 (42.9) | <0.001 |
| Laboratory Diagnostics |  |  |  |  |  |  |
| N (CBCs) | 5,102 (100.0) | 1,526 (29.9) | 490 (9.6) | 448 (8.8) | 2,638 (51.7) |  |
| HGB [mmol/l] | 6.8 (5.5 – 7.9) | 7.1 (5.6 – 8.3) | 6.8 (5.5 – 8.4) | 6.9 (5.6 – 8.1) | 6.6 (5.5 – 7.6) | <0.001 |
| MCV [fl] | 91.0 (86.0 – 96.8) | 88.6 (84.2 – 93.5) | 90.1 (85.3 – 95.9) | 90.7 (86.3 – 96.3) | 93.0 (88.0 – 98.0) | <0.001 |
| PLT [Gpt/l] | 199.0 (123.2 – 290.0) | 194.5 (113.0 – 287.0) | 204.0 (125.2 – 292.0) | 193.5 (106.0 – 275.2) | 201.0 (131.0 – 294.0) | <0.001 |
| RBC [Tpt/l] | 3.7 (3.0 – 4.3) | 3.9 (3.1 – 4.5) | 3.7 (2.9 – 4.5) | 3.8 (3.0 – 4.4) | 3.6 (3.0 – 4.2) | <0.001 |
| WBC [Gpt/l] | 12.9 (8.2 – 18.6) | 13.1 (7.8 – 18.8) | 12.7 (8.3 – 18.0) | 12.1 (7.7 – 17.7) | 13.0 (8.5 – 18.8) | 0.059 |

<sup>1</sup>n (%); Median (IQR); <sup>2</sup>Kruskal-Wallis rank sum test; Pearsons Chi-squared test; UMLT:
University of Leipzig Medical Center (Training Set); UMLV: University of Leipzig Medical
Center (Validation Set); UMG: University Medicine Greifswald; MIMIC: Medical Information
Mart for Intensive Care IV database: Medical Information Mart for Intensive Care IV database; N
(cases): total number of cases; N (CBCs): total number of blood counts; HGB: haemoglobin;
MCV: mean corpuscular volume; PLT: platelets; RBC: red blood count; WBC: white blood count

### Comparison Versus PCT

To evaluate the performance of the CBC model in comparison to the sepsis biomarker PCT, data from 17,898 cases with 24,125 simultaneous measurements of CBC and PCT, including 425 sepsis cases with 425 measurements, could be extracted from the UMLT dataset. The AUROC of the CBC+PCT model on the subset from UMLV was 0.857 (95% CI, 0.836–0.877), while the prediction performance of the biomarker PCT alone yielded an AUROC of 0.790 (95% CI, 0.759–0.821, Figure 4). The difference between both predictors was statistically significant (p<0.001).

**Figure 4:**
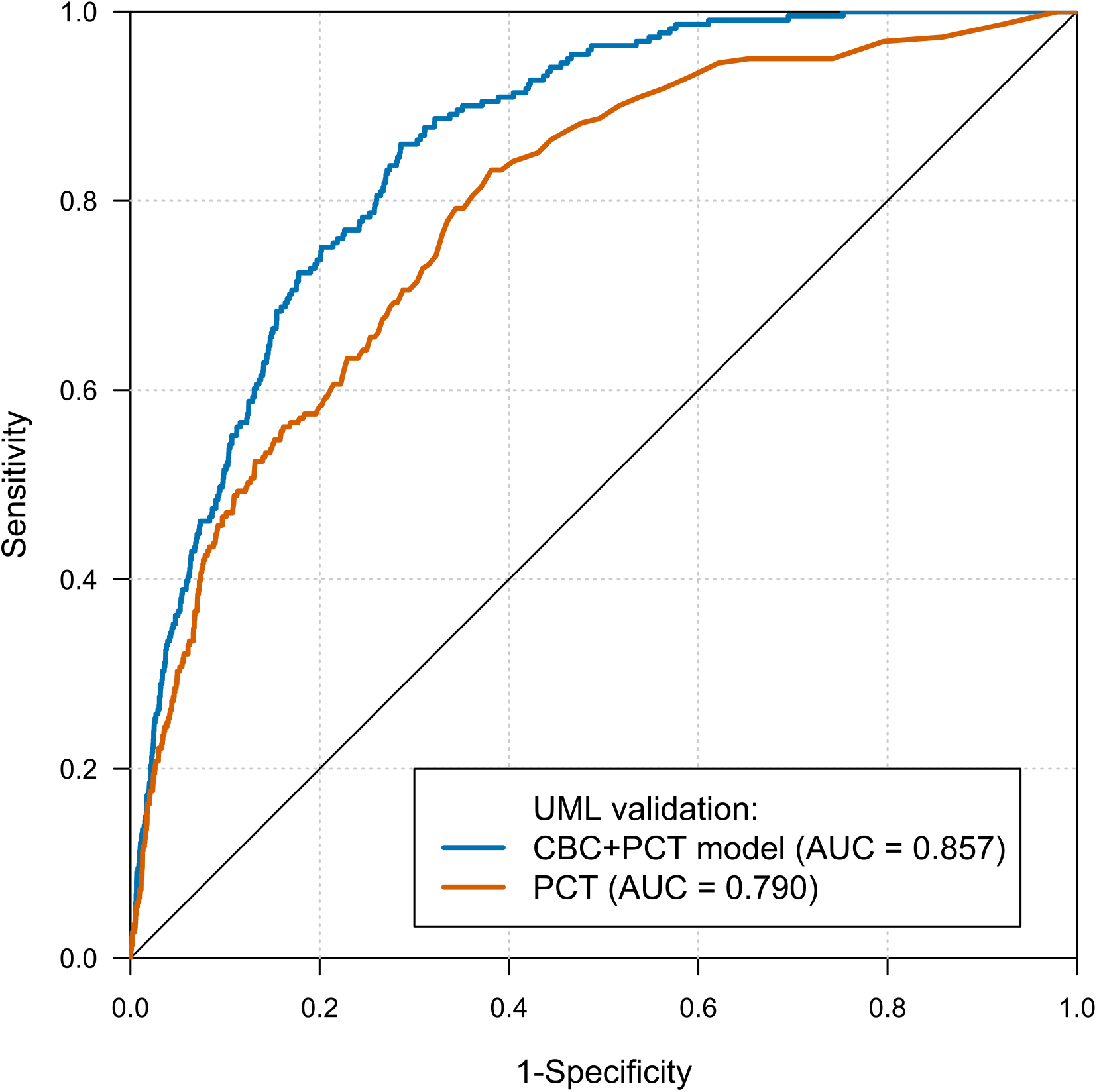
ROC curves of the comparison of the CBC+PCT model to PCT. ROC: receiver operating characteristic; AUC: area under the receiver operating characteristic; CBC: complete blood count; PCT: procalcitonin; UML: University Medicine Leipzig; The difference between both predictors is statistically significant (p<0.001, DeLong test).

## Discussion

With this study, we demonstrate the first ML-based diagnostic approach for the prediction of sepsis and need for intensive care based solely on simple blood count diagnostics. The use of broadly available CBC data in the development of clinical decision support systems (CDSS) has the potential to improve clinical outcome through earlier diagnosis.

Currently, our unique diagnostic approach can be compared with the existing literature only to a limited extent. Other studies have applied different diagnostic parameters, patient populations, ML techniques, and endpoints.^9,20,21^ Importantly, the studies have significantly preselected patient collectives (e.g., with already existing sepsis suspicion) and used divergent sepsis definitions.^22^ To date, there are no published results of comparable approaches using the publicly available MIMIC-IV dataset.

Few ML-based approaches have reported an AUROC of >0.90, but all require a multitude of input features, some of which must be recorded hourly. Among these features are variables that are rarely available from EHRs, such as vital signs, blood cultures, current medication, and cormorbidities.^23,24^ Furthermore, extensive and costly laboratory measurements are not often requested outside the ICU. Even if data are available in ICUs, the corresponding algorithms are inadequate to support the treatment of non-ICU patients. Crucial for the clinical relevance of a sepsis alert system is the prediction of patients who are not yet under intensive care and have not yet been suspected of having an infection.^25^ To date, only one study that included non-ICU patients and was based solely on laboratory diagnostics has successfully predicted sepsis. Choi et al.^26^ identified five laboratory measurements (albumin, mean platelet volume, total protein, blood urea nitrogen, and alkaline phosphatase) as the optimal combination to distinguish septic patients and patients with fever without sepsis, achieving, without external validation, an AUROC of 0.83. However, the study did not differentiate ICU and non-ICU patients, and this extensive laboratory diagnostic is rarely requested simultaneously in standard medical care in Europe. Consequently, for optimal applicability, we limited ourselves to the commonly available CBC without differentiation of leucocytes and were able to show a precise discrimination of patients with sepsis from an unselected non-ICU patient cohort (AUROC 0.872). Additionally, we are not aware of any other ML model, based on either ICU or non-ICU data, which can predict sepsis from routine laboratory diagnostics, and that has been externally validated.

Our external validations proved the robustness of our approach, with AUROCs of 0.845 and 0.805, despite significant differences in the clinical and sociodemographic characteristics of the cohorts. A recently published independent validation of the most widely implemented early warning system for sepsis in U.S. hospitals, the Epic Sepsis Model (based on demographic, vital sign, laboratory, medication, procedural, and comorbidity variables) showed a rather poor performance with an AUROC of 0.63 and thus a high risk of alert fatigue.^27^ This finding underlines the importance of external validation and reveals a major lack of research in the field of ML models.

Another important aspect of the alert system is the prediction time prior to sepsis onset. Early warnings can be achieved if a model strongly weights the “baseline risk” of developing sepsis. However, to achieve the optimal clinical impact and prevent alarm fatigue, the model must warn close to the sepsis onset. We therefore optimised our model for temporal dynamics and were able to achieve a significant discriminatory power in the period 6-48 h before ICU admission. Only a few studies addressed this temporal dynamic before. Using only CBCs, our model is superior to these, and only outperformed by models that include complex clinical data directly before sepsis onset.^9,21^ There is no study that has systematically addressed AUROCs from baseline (e.g., weeks) as well as close to sepsis onset. Therefore, the proportion of “baseline risk” in the previously published models cannot be estimated.^9,21^ In our view this is of crucial importance, as only small differences between AUROCs at sepsis onset and baseline mean a large number of alarms without temporal context to sepsis onset. Future alerts from the same patient may not be seriously considered by the treating physicians and alert fatigue could result.

PCT is considered the best-established sepsis biomarker and is used in practice as laboratory confirmation of suspected sepsis. However, its predictive performance is not sufficient to be recommended for the diagnosis of sepsis by clinical guidelines.^4^ To achieve an increased diagnostic value the presented CBC model could be combined with PCT. The resulting CBC+PCT model significantly outperforms PCT in our data set. This underlines the great potential for further development by adding more diagnostic biomarkers and will be subject of future research. However, in the interest of broad applicability and given the unexpected high predictive power of the CBC, we decided to keep the prediction model simple without addition of further diagnostic parameters.

Our study has limitations. We carried out a retrospective trial based on ICD-10 coded diagnoses. ICD-10 coding strategies have an impact on the incidence rate of sepsis and may explain differences between our training and validation sites.^28^ However, for our data extraction, we used only explicit sepsis-related ICD-10 codes and performed validations (see supplement). According to the current literature, the true incidence of sepsis is underestimated by 1.4-to 3-fold.^5,28^ For this reason, it is plausible to assume that some patients were correctly predicted by our model but not coded. The alteration of the sepsis definition during our observation time may further influence the results.^29^ The high number of patients with only mild diseases may positively influence the AUROC. Therefore, we performed cohort analyses and could demonstrate a comparable performance (see supplement). Furthermore, we did not comprehensively evaluate the chosen ML algorithm, RUSBoost. A smaller or simpler model might provide similar results and other learning algorithms might achieve a better performance.^9^ However, a systematic comparison of different machine learning approaches was outside the scope of this study. Finally, due to our selection process, the application in (post-)surgical patients can only be assessed to a limited extent.

In conclusion, we have demonstrated that CBCs contain sufficient information that, when combined with ML, facilitates early and reliable prediction of patients at risk of ICU admission due to sepsis. Our approach showed high robustness in the validations and can be calculated with any CBC diagnostic at no additional cost. CBC diagnostics are widely available, easy to perform, highly standardised, generally well digitised and have a short turn-around time. Included in a CDSS, our CBC model could reduce the time needed to detect patients with sepsis who would benefit from ICU admission. This would allow a definitive therapy to be initiated much earlier and has the potential to significantly improve clinical outcome. We are currently in the process of initiating a prospective evaluation of our CBC model as part of our laboratory based CDSS (www.ampel.care).^30^

## Data Availability

All data and code are published under the Creative Commons Attribution 4.0 International (CC BY 4.0) Public License.

https://github.com/ampel-leipzig/sbcdata

https://github.com/ampel-leipzig/sbcmodel

## Data Availability

Datasets from the UML and UMG and a conversion function for the MIMIC-IV data are available as an R package at https://github.com/ampel-leipzig/sbcdata. The MATLAB scripts are also available at https://github.com/ampel-leipzig/sbcmodel. All data and code are published under the Creative Commons Attribution 4.0 International (CC BY 4.0) Public License.

## Author Contributions

Funding was acquired by TK. The study was conceptualised by SG, DS, MF, MS, PA, TK. The Data was acquired by MF. The data was analysed by SG, DS, PA. Software was written by SG, DS, PA. Original drafts of the manuscript were written by SG, DS, PA, TK. Drafts were edited and reviewed by CL, LH, MF, MG, MN, BI, MS. Visualisations were produced by SG, MS, PA. The model was validated with a randomly selected prospective patient cohort by LH. The project was supervised by SG, TK.

## Declaration of Interests

### No competing interests

The study was supported by the AMPEL project (www.ampel.care). This project is co-financed through public funds according to the budget decided by the Saxon State Parliament under the RL eHealthSax 2017/18 grant number 100331796. The funder provided support in the form of salaries for authors PA, MS, LH, and SG. The author DS was funded by the POLAR_MI project of the German Federal Ministry of Education and Research under grant number 01ZZ1910[A-Z]. Both funders did not have any additional role in the study design, data collection and analysis, decision to publish, or preparation of the manuscript. All authors have completed the ICMJE uniform disclosure form at http://www.icmje.org/disclosure-of-interest/ and declare: no support from any organisation for the submitted work; no financial relationships with any organisations that might have an interest in the submitted work in the previous three years; no other relationships or activities that could appear to have influenced the submitted work.

## Acknowledgments

We thank the whole AMPEL team for their support, including Mark Wernsdorfer for his preparatory work and Stefan Bollmann and Thomas Hildebrandt for retrieving laboratory and administrative data at University Medicine Greifswald and Manuela Gerber for selective individual chart review at University Medicine Greifswald.

## Supplementary Material

### Validation of sepsis label based on ICD-10 Codes

Since it was not possible for us to separate sepsis from ICU admission, we compared the validation set based on ICD-10 coded administration data with data prospectively collected as part of our quality improvement initiative (10.1097/CCM.0000000000002069) at UMG. The dataset of the initiative contains only patients with severe sepsis or septic shock. We found that patients from the medical ICU (only data for 2015 and 2016) and surgical ICU (2015-2020) that are present in both databases fulfil the sepsis definition at the time of ICU admission (difference of ICU admission and sepsis onset according to the sepsis definition, medical patients: -0.6 h [-1.4,0.6], Median [25%,75% quartile]); surgical patients: 0.8 hours [-0.3,6.9], Median [25%,75% quartile]).

Given the size of our data set, it was not possible for us to check all cases individually. However, we manually examined sample subsets at UML. In 60 random cases identified as “true positives” we detected 9 cases (15%) with sepsis onset >7d after ICU admission. In 89 random cases identified as “true positives” we found 2 cases (2%) that were labeled with sepsis ICD-10 codes without this being plausibly supported by the patient file. A random sample of 75 “false positives” revealed that 4 cases (5%) had a diagnosis of sepsis in the discharge letter but were not coded with ICD. This is significantly higher than would be expected given the sepsis prevalence in the data set (0.13%). Interestingly, symptomatic infection was identified in 40 cases (53%).

### Sepsis Related ICD-10 Codes

Cases coded with one of the following International Statistical Classification of Diseases and Related Health Problems - Version 10 (ICD-10) codes were labelled as *sepsis*:

- A02.1
- A20.7
- A22.7
- A24.1
- A26.7
- A32.7
- A39.2, A39.3, A39.4
- A40.0, A40.1, A40.2, A40.3, A40.8, A40.9
- A41.0, A41.1, A41.2, A41.3, A41.4, A41.51, A41.52, A41.58, A41.8, A41.9
- A42.7
- B37.7
- R57.2
- R65.1

For the Medical Information Mart for Intensive Care IV database (MIMIC-IV) dataset sepsis cases were labelled accordingly, including additional ICD-10 and ICD-9 codes:

- A41.01, A41.02
- A41.53, A41.59, A41.81, A41.89
- R65.2, R65.20, R65.21
- 785.52
- 995.91, 995.92

### Data Processing and Model Development

We tuned the hyperparameters, including number of trees, maximum tree depth and learning rate in a 5-fold cross-validation to maximize the area under the receiver operating characteristic (AUROC). For internal validation (data not shown) the optimized hyperparameters were applied in a second 5-fold cross validation and the mean AUROC of the 5 sub models was calculated. The final CBC model was then trained on the whole UMLT training dataset. The same procedure was followed for training the modified CBC+PCT model on preselected data points including PCT measurements. All validations were performed on data sets that were unknown for model training. The high-level diagram of this workflow is displayed in figure S1.

Following final hyperparameters of the CBC model were used:

- Number of trees: 700
- Maximum split count (tree depth): 40
- Learning rate: 1

### Laboratory Diagnostics

The blood was collected in EDTA tubes (S-Monovette®, Sarstedt, Nürnbrecht, Germany for University of Leipzig Medical Centre (UML) and BD-Vacutainer®, Becton, Dickinson and Company, Franklin Lakes, USA for University Medicine Greifswald (UMG)). At UML the complete blood counts (CBCs) were analysed within 4 hours at the laboratory on a Sysmex XN-10 in an XN-9000 or XN-9100 configuration. The Institute of Clinical Chemistry and Laboratory Medicine (IKCL) at the UMG used the same Sysmex device to analyse the CBC.

**Figure S1:**
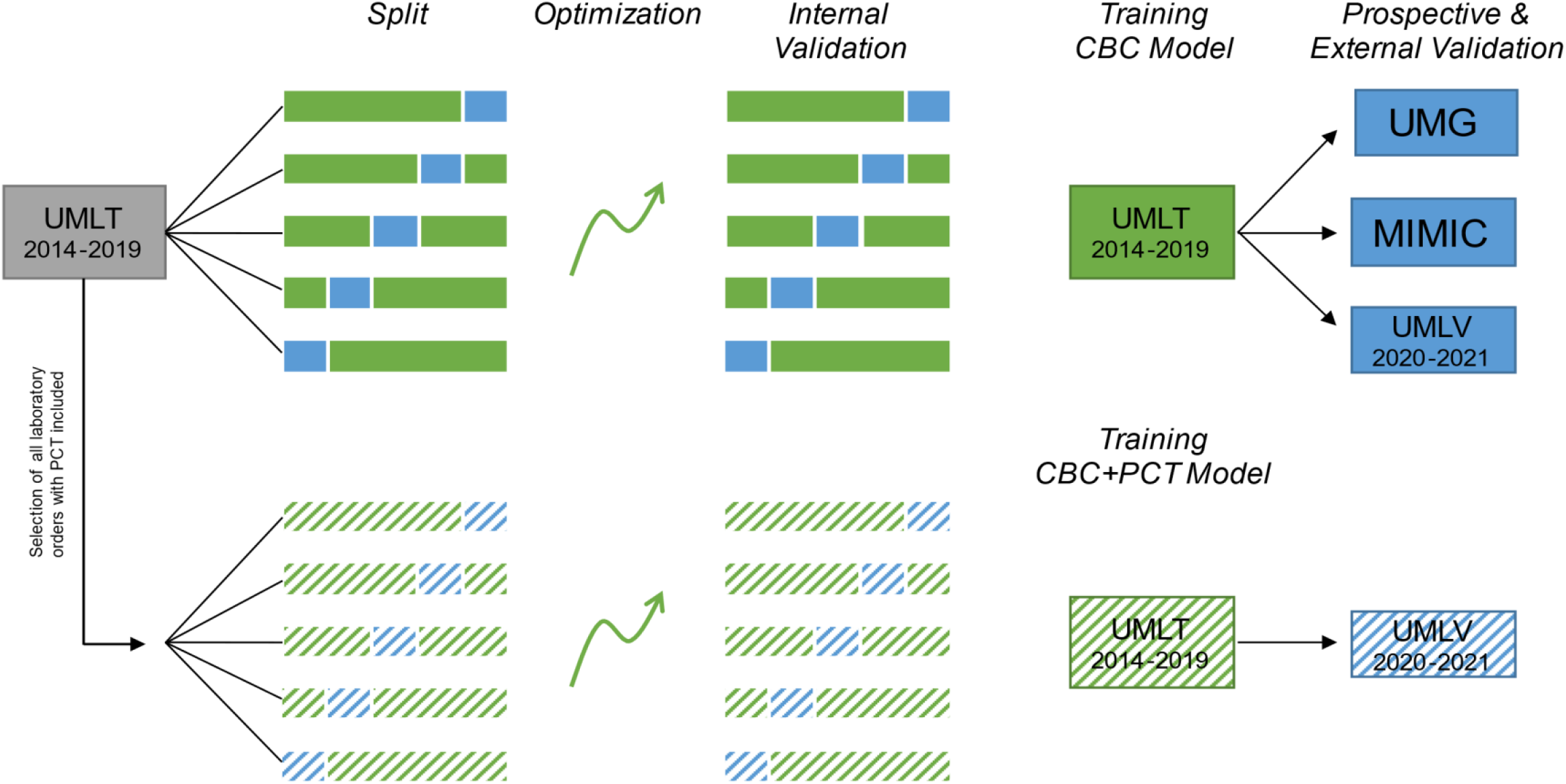
High level workflow of the model development. UMLT: University of Leipzig Medical Centre (Training Set); UMLV: University of Leipzig Medical Centre (Validation Set); UMG: University Medicine Greifswald; MIMIC: Medical Information Mart for Intensive Care IV database; CBC: complete blood count; PCT: procalcitonin

**Figure S2:**
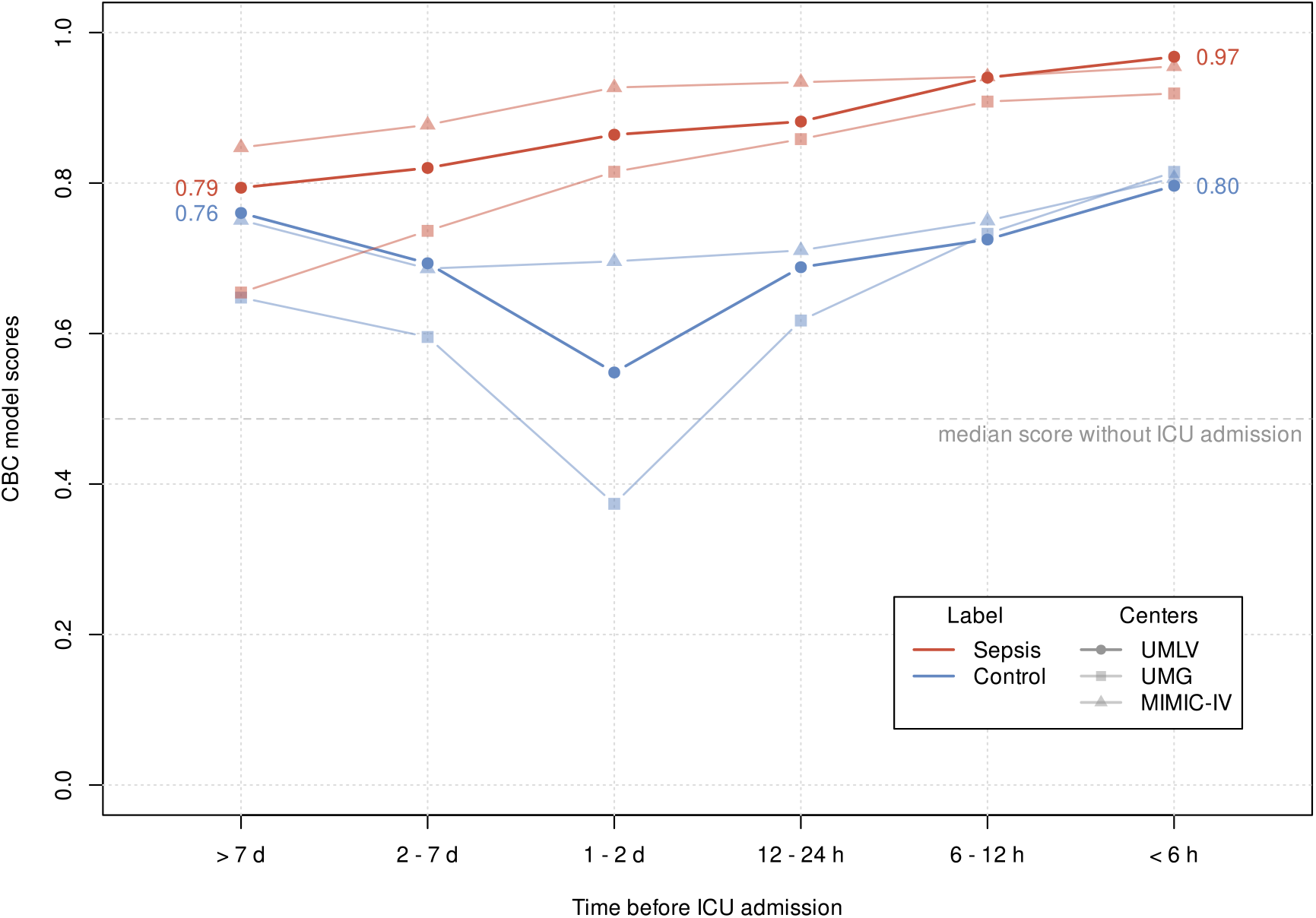
Median CBC model scores of patients with ICU admission. CBC: complete blood count; ICU: intensive care unit, UMLV: University of Leipzig Medical Centre (Validation Set); UMG: University Medicine Greifswald; MIMIC-IV: Medical Information Mart for Intensive Care IV database; Displayed are the score values of the CBC model over time comparing sepsis cases and cases with ICU admission but without sepsis diagnosis. In all three validation data sets, the largest difference is detected between 6 and 48 hours before ICU admission.

**Figure S3:**
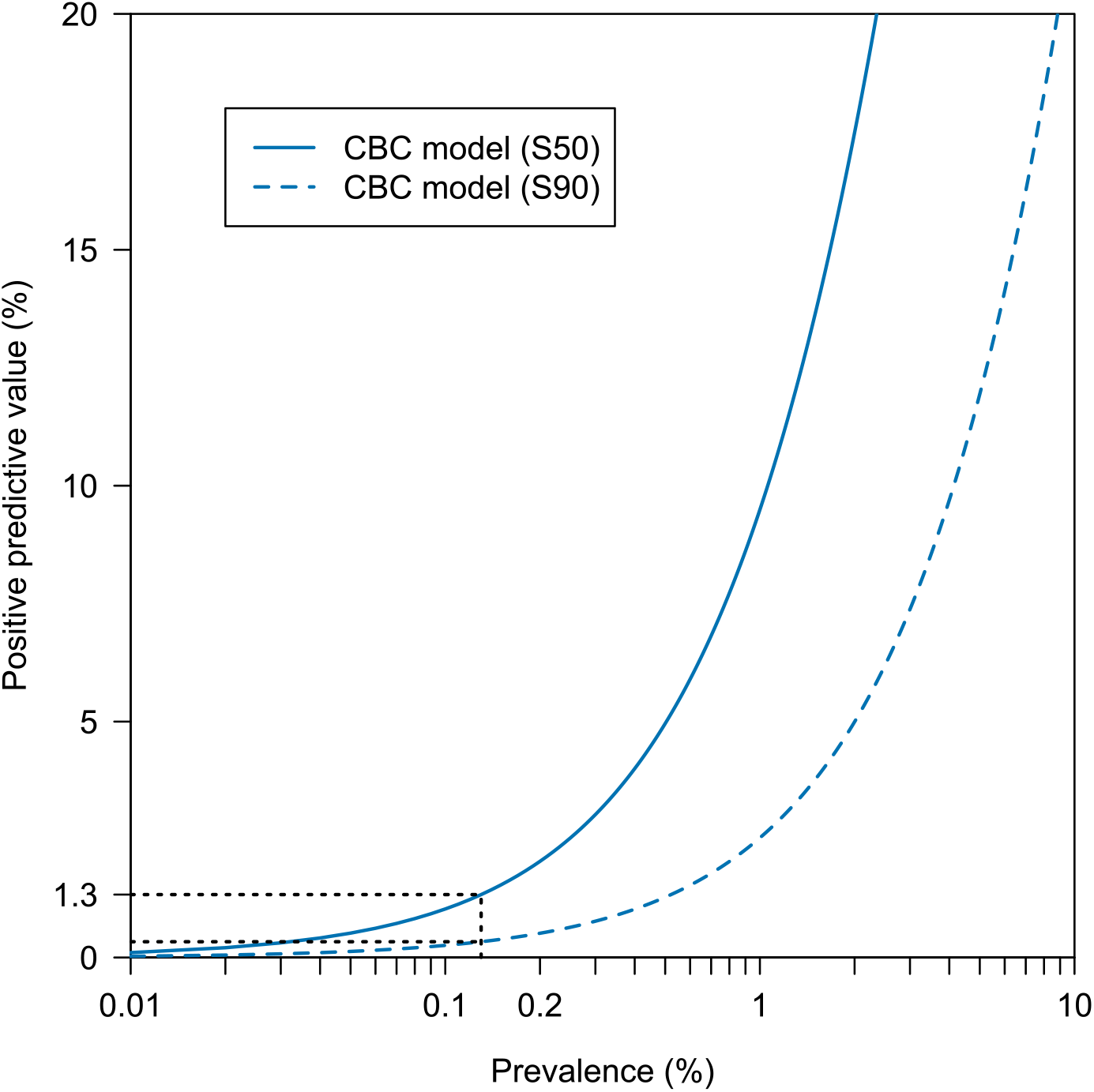
The positive predictive value in dependence of prevalence. CBC: complete blood count; correlation of prevalence and positive predictive value by different cut-offs of the CBC model with sensitivity of 50% (S50) and 90% (S90) based on UMLV data.

**Figure S4:**
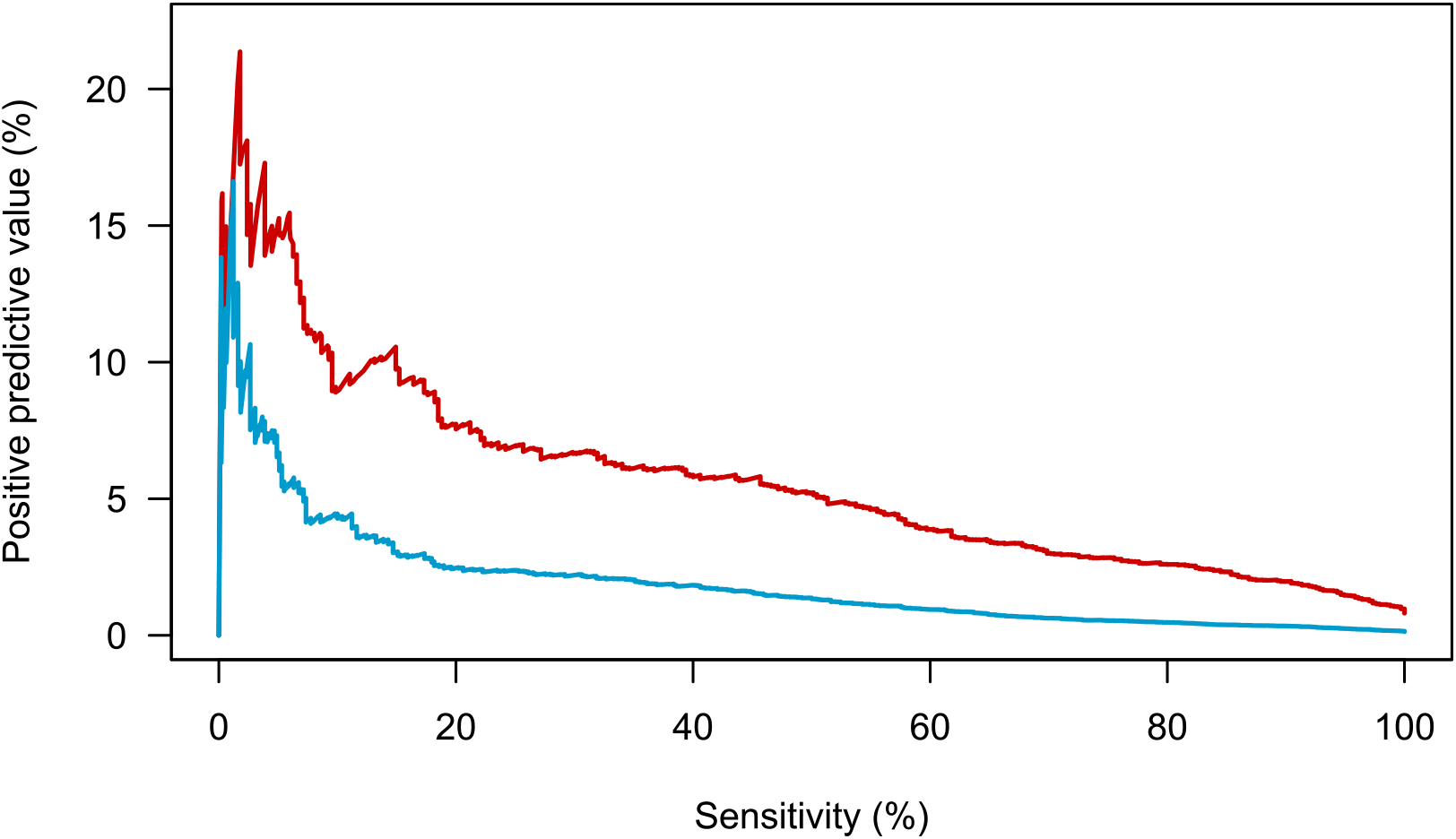
The positive predictive value in dependence of sensitivity. Correlation of sensitivity and positive predictive for two cohorts with different prevalences based on UMLV data. Graphs were derived from all patients with a sepsis prevalence of 0.13% (blue) and the emergency department cohort with a prevalence of 0.81% (red).

**Figure S5:**
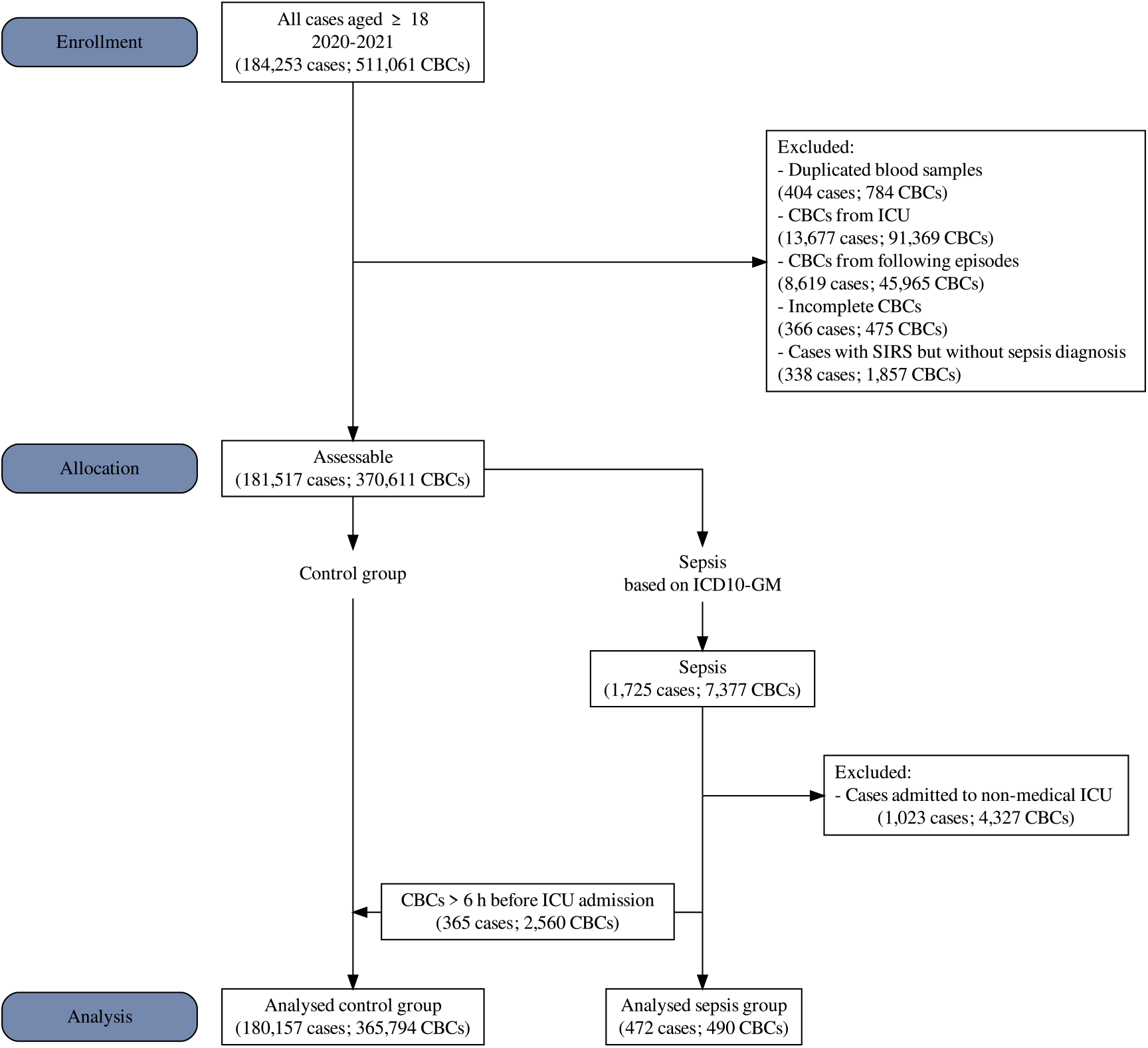
The flowchart of the inclusion criteria and the size of the validation dataset from UMLV (2020-2021). UMLV: University of Leipzig Medical Centre (Validation Set); CBC: complete blood count; ICU: intensive care unit; SIRS: systemic inflammatory response syndrome; ICD-10-GM: International Statistical Classification of Diseases and Related Health Problems Version 10 - German Modification.

**Figure S6:**
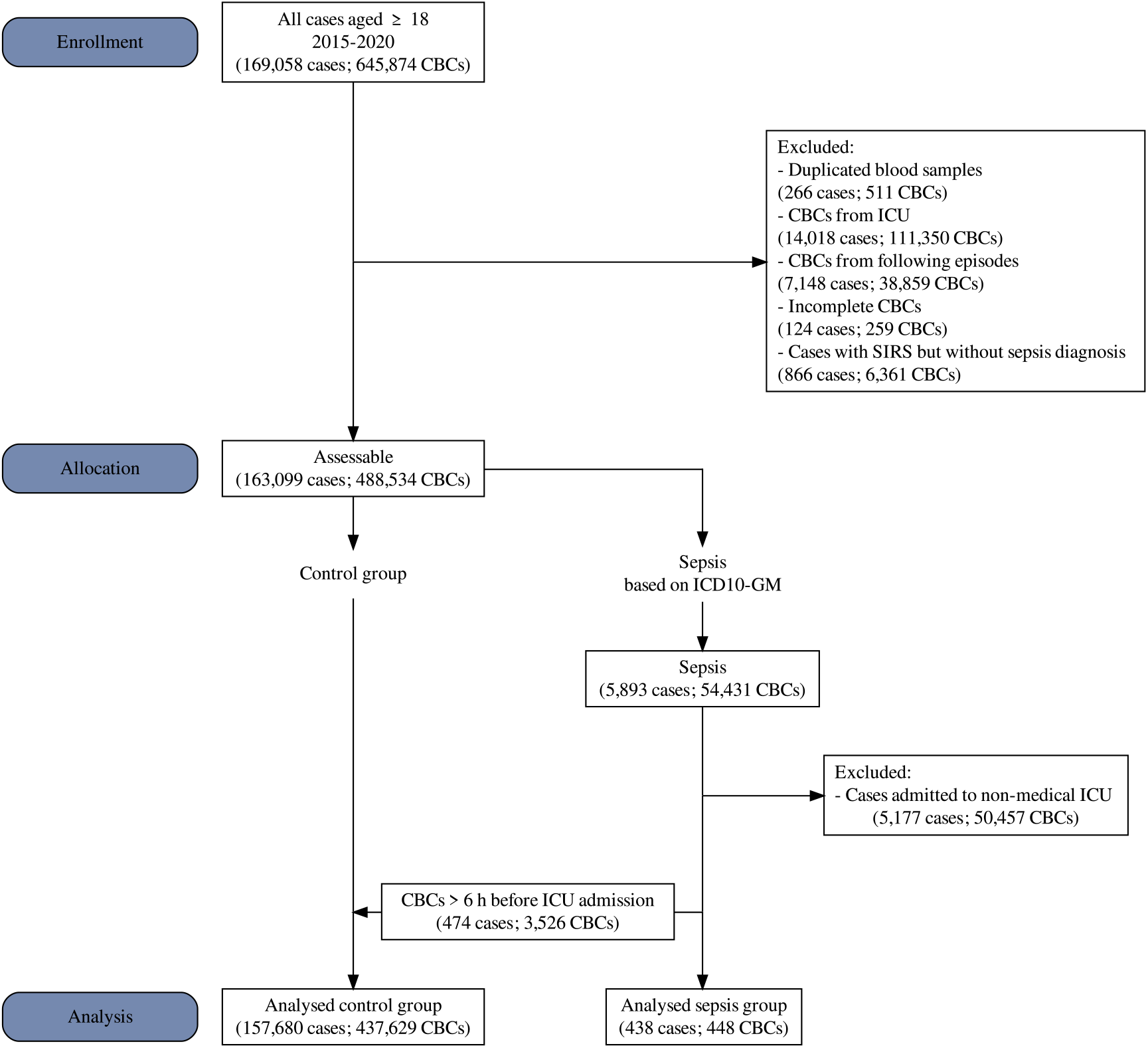
The flowchart of the inclusion criteria and the size of the validation dataset from UMG. UMG: University Medicine Greifswald; CBC: complete blood count; ICU: intensive care unit; SIRS: systemic inflammatory response syndrome; ICD-10-GM: International Statistical Classification of Diseases and Related Health Problems Version 10 - German Modification

**Figure S7:**
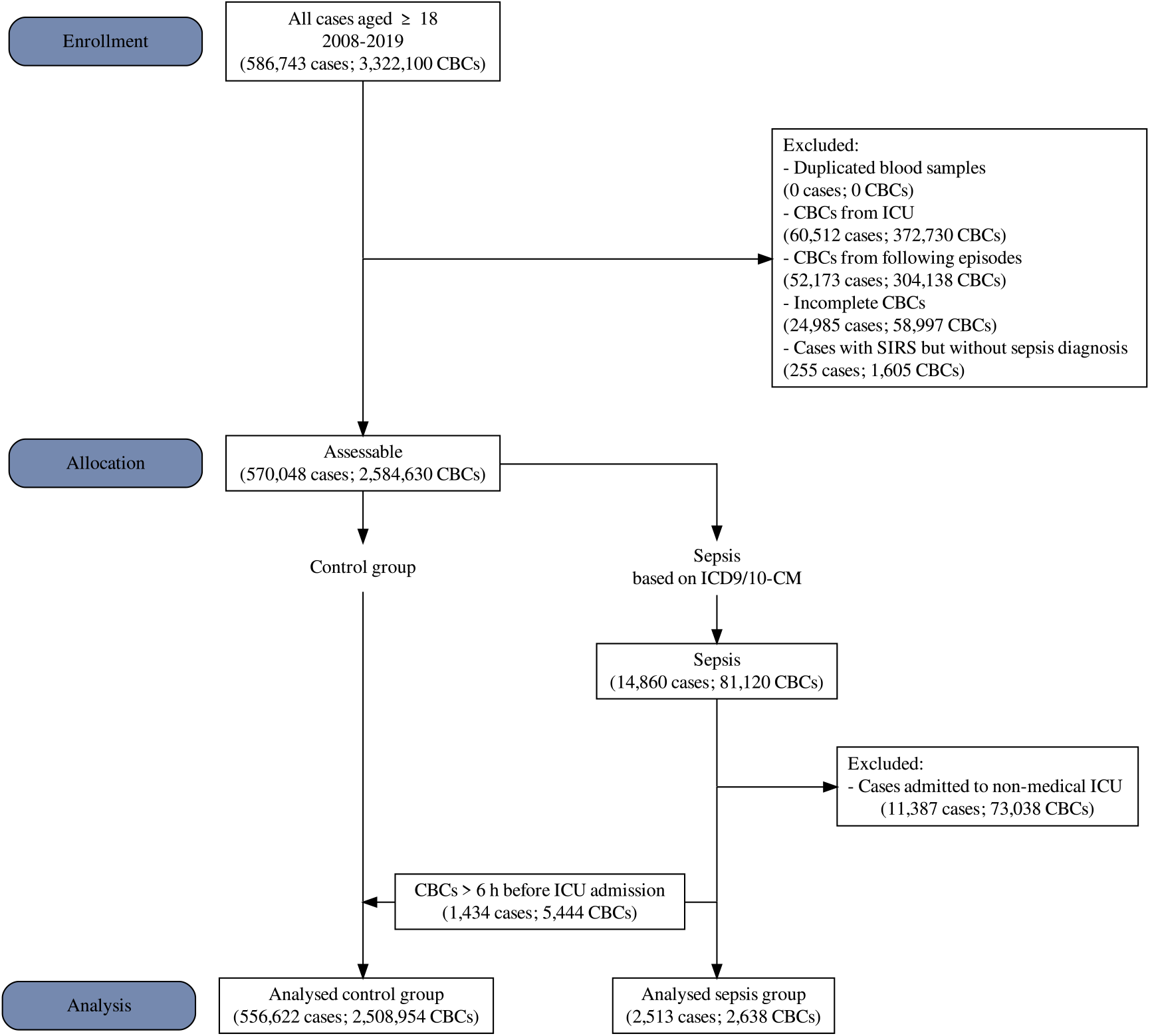
The flowchart of the inclusion criteria and the size of the validation dataset from MIMIC-IV. MIMIC-IV: Medical Information Mart for Intensive Care IV database; CBC: complete blood count; ICU: intensive care unit; SIRS: systemic inflammatory response syndrome; ICD9/10-CM: International Statistical Classification of Diseases and Related Health Problems Version 9 and 10

**Table S1.1:** Cohort analysis of all patients at the three centres UML, UMG, and MIMIC-IV.

| Cohorts | UMLV |  |  |  |  | UMG |  |  |  |  | MIMIC-IV |  |  |  |  |
| --- | --- | --- | --- | --- | --- | --- | --- | --- | --- | --- | --- | --- | --- | --- | --- |
|  | N | PRE<sup>1</sup> | PPV<sup>1</sup> | NPV<sup>1</sup> | AUC | N | PRE<sup>1</sup> | PPV<sup>1</sup> | NPV<sup>1</sup> | AUC | N | PRE<sup>1</sup> | PPV<sup>1</sup> | NPV<sup>1</sup> | AUC |
| Overall | 490 | 0.13 | 1.37<br>(1.20-1.55) | 99.93<br>(99.92-99.94) | 0.872<br>(0.857-0.887) | 448 | 0.10 | 0.38<br>(0.33-0.44) | 99.94<br>(99.93-99.95) | 0.805<br>(0.787-0.824) | 263<br>8 | 0.11 | 0.78<br>(0.74-0.82) | 99.94<br>(99.94-99.95) | 0.845<br>(0.837-0.852) |
| Age < 60 | 134 | 0.08 | 0.56<br>(0.43-0.71) | 99.96<br>(99.95-99.97) | 0.888<br>(0.862-0.914) | 126 | 0.08 | 0.32<br>(0.25-0.41) | 99.95<br>(99.94-99.96) | 0.839<br>(0.812-0.866) | 753 | 0.07 | 0.55<br>(0.49-0.60) | 99.96<br>(99.96-99.97) | 0.865<br>(0.852-0.878) |
| Age 60-80 | 256 | 0.18 | 1.82<br>(1.52-2.16) | 99.91<br>(99.89-99.92) | 0.854<br>(0.830-0.879) | 234 | 0.12 | 0.42<br>(0.35-0.50) | 99.93<br>(99.92-99.94) | 0.801<br>(0.775-0.827) | 117<br>5 | 0.12 | 0.78<br>(0.72-0.85) | 99.94<br>(99.93-99.94) | 0.825<br>(0.813-0.838) |
| Age > 80 | 100 | 0.22 | 1.27<br>(0.94-1.67) | 99.88<br>(99.84-99.91) | 0.829<br>(0.789-0.869) | 88 | 0.10 | 0.43<br>(0.31-0.58) | 99.94<br>(99.92-99.96) | 0.790<br>(0.742-0.837) | 710 | 0.18 | 1.08<br>(0.97-1.20) | 99.90<br>(99.89-99.91) | 0.809<br>(0.792-0.827) |

<sup>1</sup> % (95% CI); UMLV: University of Leipzig Medical Centre (Validation Set 2020-2021); UMG: University Medicine Greifswald; MIMIC-IV: Medical Information Mart for Intensive Care IV database; N: total number of complete blood counts with sepsis label; PRE: prevalence; PPV: positive predictive value; NPV: negative predictive value; AUC: area under the receiver operating characteristic; All values are calculated with a sensitivity of 50%.

**Table S1.2:**
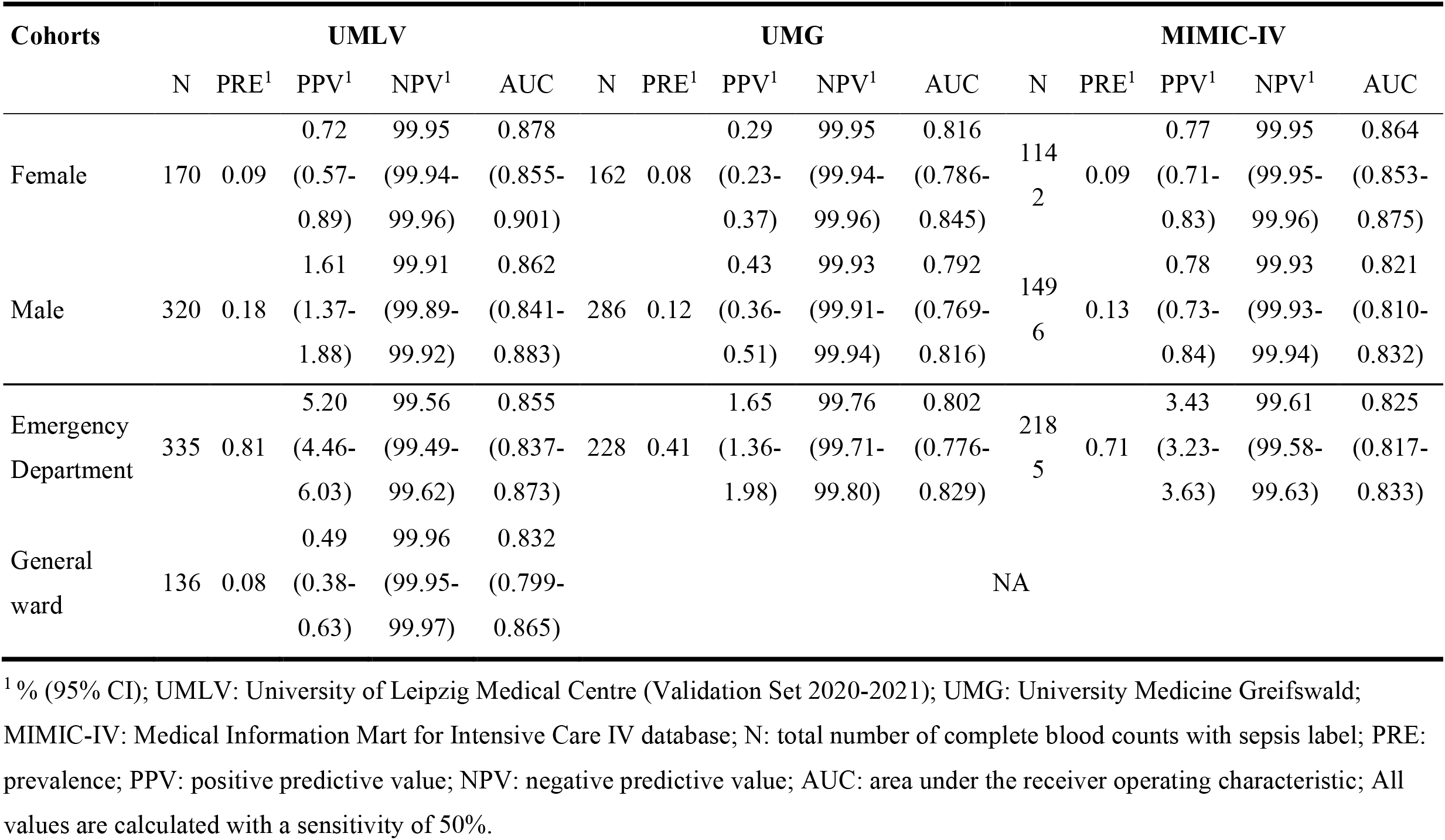
Cohort analysis of all patients at the three centres UML, UMG, and MIMIC-IV.

**Table S2:** Baseline characteristics of all patients at UML (Training Set 2014-2019).

| Characteristic | Overall<sup>1</sup> | Control<sup>1</sup> | Sepsis<sup>1</sup> | p-value<sup>2</sup> |
| --- | --- | --- | --- | --- |
| General |  |  |  |  |
| N (cases) | 528,526 (100.0) | 527,038 (99.7) | 1,488 (0.3) |  |
| Age | 57 (38 – 71) | 57 (38 – 71) | 67 (57 – 76) | <0.001 |
| Male | 252,905 (47.9) | 251,967 (47.8) | 938 (63.0) | <0.001 |
| Female | 275,621 (52.1) | 275,071 (52.2) | 550 (37.0) | <0.001 |
| Laboratory Diagnostics |  |  |  |  |
| N (CBCs) | 1,015,074 (100.0) | 1,013,548 (99.8) | 1,526 (0.2) | <0.001 |
| HGB [mmol/l] | 7.8 (6.5 – 8.7) | 7.8 (6.5 – 8.7) | 7.1 (5.6 – 8.3) | <0.001 |
| MCV [fl] | 87.7 (84.3 – 91.2) | 87.7 (84.3 – 91.2) | 88.6 (84.2 – 93.5) | <0.001 |
| PLT [Gpt/l] | 236.0 (180.0 – 296.0) | 236.0 (181.0 – 296.0) | 194.5 (113.0 – 287.0) | <0.001 |
| RBC [Tpt/l] | 4.2 (3.6 – 4.7) | 4.2 (3.6 – 4.7) | 3.9 (3.1 – 4.5) | <0.001 |
| WBC [Gpt/l] | 7.4 (5.6 – 9.8) | 7.4 (5.6 – 9.8) | 13.1 (7.8 – 18.8) | <0.001 |

<sup>1</sup>n (%); Median (IQR); <sup>2</sup>Wilcoxon rank sum test; Pearsons Chi-squared test; UML: University of Leipzig Medical Centre; N (cases): total number of cases; N (CBCs): total number of complete blood counts; HGB: haemoglobin; MCV: mean corpuscular volume; PLT: platelets; RBC: red blood count; WBC: white blood count

**Table S3:** Baseline characteristics of all patients at UML (Validation Set 2020-2021).

| Characteristic | Overall<sup>1</sup> | Control<sup>1</sup> | Sepsis<sup>1</sup> | p-value<sup>2</sup> |
| --- | --- | --- | --- | --- |
| General |  |  |  |  |
| N (cases) | 180,629 (100.0) | 180,157 (99.7) | 472 (0.3) |  |
| Age | 58 (39 – 71) | 58 (39 – 70) | 68 (59 – 78) | <0.001 |
| Male | 86,851 (48.1) | 86,541 (48.0) | 310 (65.7) | <0.001 |
| Female | 93,778 (51.9) | 93,616 (52.0) | 162 (34.3) | <0.001 |
| Laboratory Diagnostics |  |  |  |  |
| N (CBCs) | 366,284 (100.0) | 365,794 (99.9) | 490 (0.1) | <0.001 |
| HGB [mmol/l] | 7.6 (6.3 – 8.6) | 7.6 (6.3 – 8.6) | 6.8 (5.5 – 8.4) | <0.001 |
| MCV [fl] | 88.7 (85.2 – 92.4) | 88.7 (85.2 – 92.4) | 90.1 (85.3 – 95.9) | <0.001 |
| PLT [Gpt/l] | 235.0 (175.0 – 297.0) | 235.0 (176.0 – 297.0) | 204.0 (125.2 – 292.0) | <0.001 |
| RBC [Tpt/l] | 4.1 (3.4 – 4.6) | 4.1 (3.4 – 4.6) | 3.7 (2.9 – 4.5) | <0.001 |
| WBC [Gpt/l] | 7.2 (5.3 – 9.5) | 7.2 (5.3 – 9.5) | 12.7 (8.3 – 18.0) | <0.001 |

<sup>1</sup>n (%); Median (IQR); <sup>2</sup>Wilcoxon rank sum test; Pearsons Chi-squared test; UML: University of Leipzig Medical Centre; N (cases): total number of cases; N (CBCs): total number of complete blood counts; HGB: haemoglobin; MCV: mean corpuscular volume; PLT: platelets; RBC: red blood count; WBC: white blood count

**Table S4:** Baseline characteristics of all patients at UMG.

| Characteristic | Overall<sup>1</sup> | Control<sup>1</sup> | Sepsis<sup>1</sup> | p-value<sup>2</sup> |
| --- | --- | --- | --- | --- |
| General |  |  |  |  |
| N (cases) | 158,118 (100.0) | 157,680 (99.7) | 438 (0.3) |  |
| Age | 63 (50 – 76) | 63 (50 – 76) | 69 (58 – 78) | <0.001 |
| Male | 79,162 (50.1) | 78,884 (50.0) | 278 (63.5) | <0.001 |
| Female | 78,956 (49.9) | 78,796 (50.0) | 160 (36.5) | <0.001 |
| Laboratory Diagnostics |  |  |  |  |
| N (CBCs) | 438,077 (100.0) | 437,629 (99.9) | 448 (0.1) | <0.001 |
| HGB [mmol/l] | 7.5 (6.2 – 8.5) | 7.5 (6.2 – 8.5) | 6.9 (5.6 – 8.1) | <0.001 |
| MCV [fl] | 90.3 (86.8 – 94.1) | 90.3 (86.8 – 94.1) | 90.7 (86.3 – 96.3) | 0.053 |
| PLT [Gpt/l] | 225.0 (173.0 – 285.0) | 225.0 (173.0 – 285.0) | 193.5 (106.0 – 275.2) | <0.001 |
| RBC [Tpt/l] | 4.1 (3.4 – 4.6) | 4.1 (3.4 – 4.6) | 3.8 (3.0 – 4.4) | <0.001 |
| WBC [Gpt/l] | 8.1 (6.2 – 10.7) | 8.1 (6.2 – 10.7) | 12.1 (7.7 – 17.7) | <0.001 |

<sup>1</sup>n (%); Median (IQR); <sup>2</sup>Wilcoxon rank sum test; Pearsons Chi-squared test; UMG: University Medicine Greifswald; N (cases): total number of cases; N (CBCs): total number of complete blood counts; HGB: haemoglobin; MCV: mean corpuscular volume; PLT: platelets; RBC: red blood count; WBC: white blood count

**Table S5:** Baseline characteristics of all patients from MIMIC-IV.

| Characteristic | Overall<sup>1</sup> | Control<sup>1</sup> | Sepsis<sup>1</sup> | p-value<sup>2</sup> |
| --- | --- | --- | --- | --- |
| General |  |  |  |  |
| N (cases) | 559,135 (100.0) | 556,622 (99.6) | 2,513 (0.4) |  |
| Age | 58 (41 – 72) | 58 (41 – 72) | 69 (58 – 81) | <0.001 |
| Male | 258,824 (46.3) | 257,389 (46.2) | 1,435 (57.1) | <0.001 |
| Female | 300,311 (53.7) | 299,233 (53.8) | 1,078 (42.9) | <0.001 |
| Laboratory Diagnostics |  |  |  |  |
| N (CBCs) | 2,511,592 (100.0) | 2,508,954 (99.9) | 2,638 (0.1) | <0.001 |
| HGB [mmol/l] | 7.2 (6.1 – 8.1) | 7.2 (6.1 – 8.1) | 6.6 (5.5 – 7.6) | <0.001 |
| MCV [fl] | 91.0 (86.0 – 95.0) | 91.0 (86.0 – 95.0) | 93.0 (88.0 – 98.0) | <0.001 |
| PLT [Gpt/l] | 227.0 (168.0 – 292.0) | 227.0 (168.0 – 292.0) | 201.0 (131.0 – 294.0) | <0.001 |
| RBC [Tpt/l] | 3.9 (3.3 – 4.4) | 3.9 (3.3 – 4.4) | 3.6 (3.0 – 4.2) | <0.001 |
| WBC [Gpt/l] | 7.2 (5.3 – 9.6) | 7.2 (5.3 – 9.6) | 13.0 (8.5 – 18.8) | <0.001 |

<sup>1</sup>n (%); Median (IQR); <sup>2</sup>Wilcoxon rank sum test; Pearsons Chi-squared test; MIMIC-IV: Medical Information Mart for Intensive Care IV database; N (cases): total number of cases; N (CBCs): total number of complete blood counts; HGB: haemoglobin; MCV: mean corpuscular volume; PLT: platelets; RBC: red blood count; WBC: white blood count

**Table S6:** Baseline characteristics of all patients at the three centres UML, UMG, and MIMIC-IV.

| Characteristic | Overall<sup>1</sup> | UMLT<sup>1</sup> | UMLV<sup>1</sup> | UMG<sup>1</sup> | MIMIC-IV<sup>1</sup> | p-value<sup>2</sup> |
| --- | --- | --- | --- | --- | --- | --- |
| General |  |  |  |  |  |  |
| N (cases) | 1,425,178<br>(100.0) | 528,101<br>(37.1) | 180,494<br>(12.7) | 157,922<br>(11.1) | 558,661<br>(39.2) |  |
| Age | 58 (40.0 – 72) | 57 (38.0 – 71) | 58 (39.0 – 71) | 63 (50.0 – 76) | 58 (41.0 – 72) | <0.001 |
| Male | 677,016<br>(47.5) | 252,639<br>(47.8) | 86,770<br>(48.1) | 79,039<br>(50.0) | 258,568<br>(46.3) | <0.001 |
| Female | 748,162<br>(52.5) | 275,462<br>(52.2) | 93,724<br>(51.9) | 78,883<br>(50.0) | 300,093<br>(53.7) | <0.001 |
| Control | 1,421,497<br>(99.7) | 527,038<br>(99.8) | 180,157<br>(99.8) | 157,680<br>(99.8) | 556,622<br>(99.6) |  |
| Sepsis | 3,681 (0.3) | 1,063 (0.2) | 337 (0.2) | 242 (0.2) | 2,039 (0.4) |  |
| Laboratory Diagnostics |  |  |  |  |  |  |
| N (CBCs) | 4,331,027<br>(100.0) | 1,015,074<br>(23.4) | 366,284<br>(8.5) | 438,077<br>(10.1) | 2,511,592<br>(58.0) |  |
| HGB [mmol/l] | 7.4 (6.2 – 8.4) | 7.8 (6.5 – 8.7) | 7.6 (6.3 – 8.6) | 7.5 (6.2 – 8.5) | 7.2 (6.1 – 8.1) | <0.001 |
| MCV [fl] | 89.7 (85.9 – 94.0) | 87.7 (84.3 – 91.2) | 88.7 (85.2 – 92.4) | 90.3 (86.8 – 94.1) | 91.0 (86.0 – 95.0) | <0.001 |
| PLT [Gpt/l] | 229.0 (172.0 – 293.0) | 236.0 (180.0 – 296.0) | 235.0 (175.0 – 297.0) | 225.0 (173.0 – 285.0) | 227.0 (168.0 – 292.0) | <0.001 |
| RBC [Tpt/l] | 4.0 (3.4 – 4.5) | 4.2 (3.6 – 4.7) | 4.1 (3.4 – 4.6) | 4.1 (3.4 – 4.6) | 3.9 (3.3 – 4.4) | <0.001 |
| WBC [Gpt/l] | 7.3 (5.5 – 9.7) | 7.4 (5.6 – 9.8) | 7.2 (5.3 – 9.5) | 8.1 (6.2 – 10.7) | 7.2 (5.3 – 9.6) | <0.001 |

<sup>1</sup>n (%); Median (IQR); <sup>2</sup>Kruskal-Wallis rank sum test; Pearsons Chi-squared test; UMLT: University of Leipzig Medical Centre (Training Set 2014-2019); UMLV: University of Leipzig Medical Centre (Validation Set 2020-2021); UMG: University Medicine Greifswald; MIMIC-IV: Medical Information Mart for Intensive Care IV database; N (cases): total number of cases; N (CBCs): total number of complete blood counts; HGB: haemoglobin; MCV: mean corpuscular volume; PLT: platelets; RBC: red blood count; WBC: white blood count

**Table S7:** Baseline characteristics of patients included in the UML subgroup analysis for comparison with procalcitonin.

| Characteristic | Overall<sup>1</sup> | Control<sup>1</sup> | Sepsis<sup>1</sup> | p-value<sup>2</sup> |
| --- | --- | --- | --- | --- |
| General |  |  |  |  |
| N (cases) | 17,898 (100.0) | 17,473 (97.6) | 425 (2.4) |  |
| Age | 61 (51 – 70) | 61 (51 – 70) | 67 (56 – 76) | <0.001 |
| Male | 10,700 (59.8) | 10,416 (59.6) | 284 (66.8) | 0.003 |
| Female | 7,198 (40.2) | 7,057 (40.4) | 141 (33.2) | 0.003 |
| Laboratory Diagnostics |  |  |  |  |
| N (CBCs) | 24,125 (100.0) | 23,700 (98.2) | 425 (1.8) |  |
| HGB [mmol/l] | 7.3 (6.1 – 8.3) | 7.3 (6.1 – 8.3) | 7.1 (6.0 – 8.4) | 0.46 |
| MCV [fl] | 86.8 (83.1 – 90.8) | 86.8 (83.1 – 90.8) | 88.2 (84.4 – 92.9) | <0.001 |
| PCT [ng/ml] | 0.1 (0.1 – 0.3) | 0.1 (0.1 – 0.3) | 1.7 (0.4 – 9.6) | <0.001 |
| PLT [Gpt/l] | 217.0 (149.0 – 304.0) | 218.0 (149.0 – 304.0) | 199.0 (120.0 – 294.0) | <0.001 |
| RBC [Tpt/l] | 4.0 (3.4 – 4.6) | 4.0 (3.4 – 4.6) | 4.0 (3.2 – 4.5) | 0.017 |
| WBC [Gpt/l] | 7.5 (5.3 – 11.0) | 7.4 (5.2 – 10.9) | 13.1 (7.7 – 19.5) | <0.001 |

<sup>1</sup>n (%); Median (IQR); <sup>2</sup>Wilcoxon rank sum test; Pearsons Chi-squared test; University of Leipzig Medical Centre: University of Leipzig Medical Centre; N (cases): total number of cases; N (CBCs): total number of complete blood counts; HGB: haemoglobin; MCV: mean corpuscular volume; PCT: procalcitonin; PLT: platelets; RBC: red blood count; WBC: white blood count

### Predictor Importance

Predictor importance was calculated using MATLAB’s built-in function “predictorImportance” for classification ensembles created by “fitcensemble”. For every tree the amount of node risk change is determined that is associated with a feature. For this, the node risk is determined for every node, which is the product of node probability and node impurity. Then, the node risk change is the difference between the parent nodes risk and the child nodes risks divided by the number of branch nodes. Finally, the predictor importance is determined by calculating a weighted average over all trees. Accordingly, the predictor importance is the weighted average over the changes in node risk for a particular feature. For a clearer presentation of the values, we have indicated the respective share of the total for every feature.

**Figure S8:**
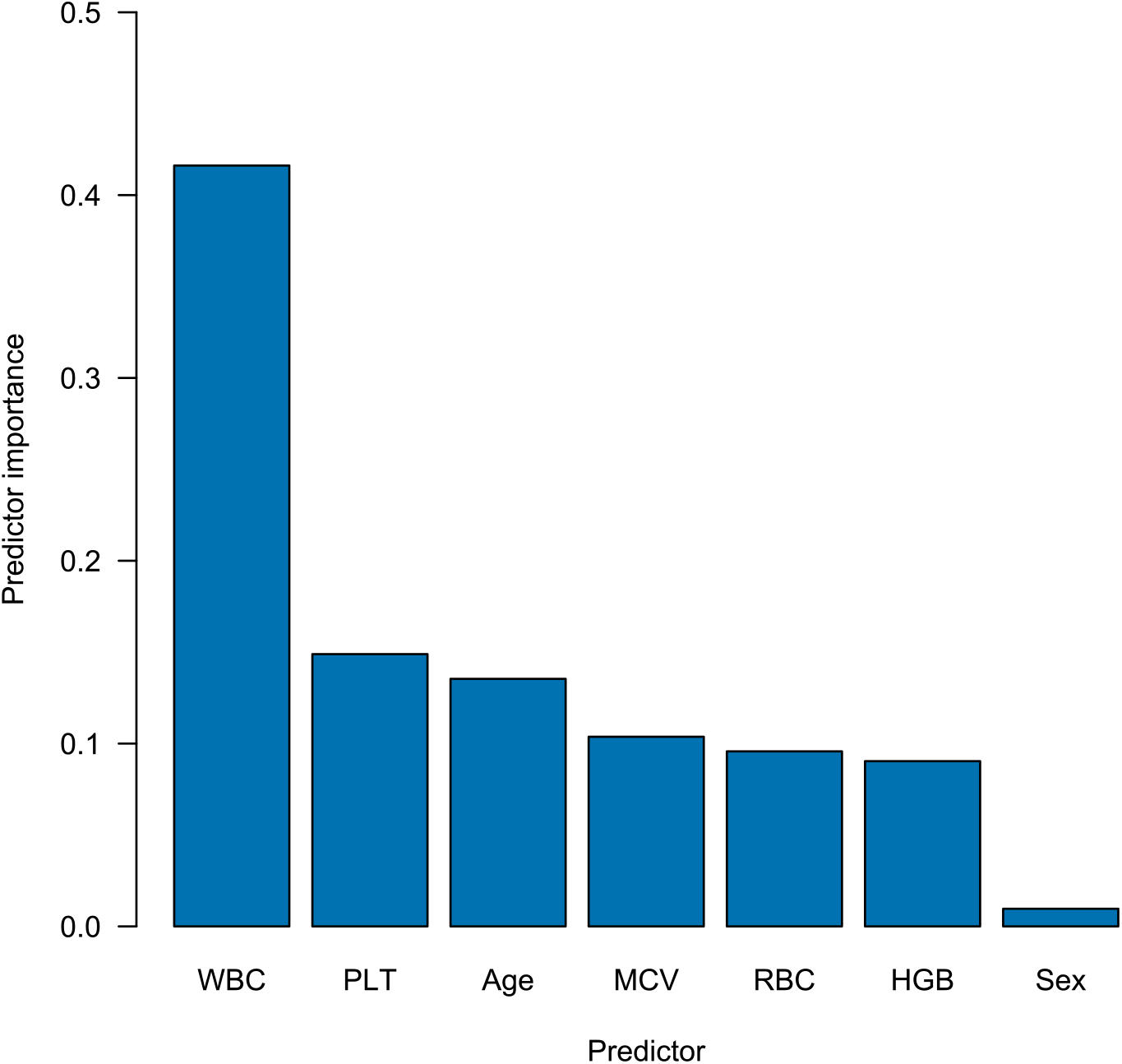
Predictor Importance. WBC: white blood count; PLT: platelets; MCV: mean corpuscular volume; RBC: red blood cells; HGB: haemoglobin

### Seed analysis

To rule out that the choice of random seed had a meaningful impact on our results, we ran the entire MATLAB pipeline for 20 different random seeds. The following seeds were used: 1969207776, 1969381777, 1969549025, 1969715444, 1969880797, 1970045850, 1970211320, 1970376985, 1970541848, 1970708391, 1970871325, 1971035844, 1971201286, 1971364957, 1971529602, 1971694292, 1971859146, 1972024133, 1972188575, 1972352809.

**Table S8:** Results of the seed analysis.

|  | AUC UMLV | AUC UMG | AUC MIMIC-IV |
| --- | --- | --- | --- |
| Maximum | 0.8770 | 0.8106 | 0.8459 |
| Minimum | 0.8720 | 0.8018 | 0.8435 |
| Standard deviation | 0.0014 | 0.0019 | 0.0006 |

AUC: area under the receiver operating characteristic; UMLV: University of Leipzig Medical Centre (Validation Set); UMG: University Medicine Greifswald; MIMIC-IV: Medical Information Mart for Intensive Care IV database

**Table S9:** Sample sizes of time windows in the validation datasets.

|  | >28d | 7d-28d | 2d-7d | 1d-2d | 12-24h | 6-12h | 0-6h |
| --- | --- | --- | --- | --- | --- | --- | --- |
| UMLV | 367 | 1062 | 624 | 205 | 115 | 190 | 490 |
| UMG | 600 | 1261 | 924 | 348 | 172 | 222 | 448 |
| MIMIC-IV | 68 | 1244 | 1784 | 762 | 595 | 994 | 2638 |

UMLV: University of Leipzig Medical Centre (Validation Set); UMG: University Medicine Greifswald; MIMIC-IV: Medical Information Mart for Intensive Care IV database

### Exploration of model behaviour

To explore the model’s behaviour for different blood count constellations, we analysed the CBC model’s decisions for different blood count values and illustrate them in the following figures. Six different cohorts were examined:

1. All patients <60y
2. All patients 60-80y
3. All patients >80y
4. Sepsis patients <60y
5. Sepsis patients 60-80y
6. Sepsis patients >80y

From every cohort, the median values were determined for the following features: Age, RBC, MCV, HGB. These values were used to generate an exemplary patient for every group. The sex was chosen to be male for all exemplary patients, since data from male patients was more prevalent in most cohorts. The two remaining features WBC and PLT were sampled in a wide range to display all clinically meaningful values. The model’s output score was plotted as a heatmap. The observed cut-offs were 0.97 (for a sensitivity of 50%) and 0.69 (for a sensitivity of 90%).

**Table S10:** Features of exemplary patients used to display model behaviour.

|  | HGB | MCV | RBC | Age |
| --- | --- | --- | --- | --- |
| All patients <60y | 8 | 87 | 4.37 | 45 |
| All patients 60-80y | 7.5 | 88.4 | 4.07 | 70 |
| All patients >80y | 7.3 | 88.7 | 3.96 | 84 |
| Sepsis patients <60 | 6.9 | 88.1 | 3.72 | 52 |
| Sepsis patients 60-80y | 7.1 | 88.6 | 3.87 | 71 |
| Sepsis patients >80y | 7.3 | 89.9 | 4.08 | 84 |

**Figure S9:**
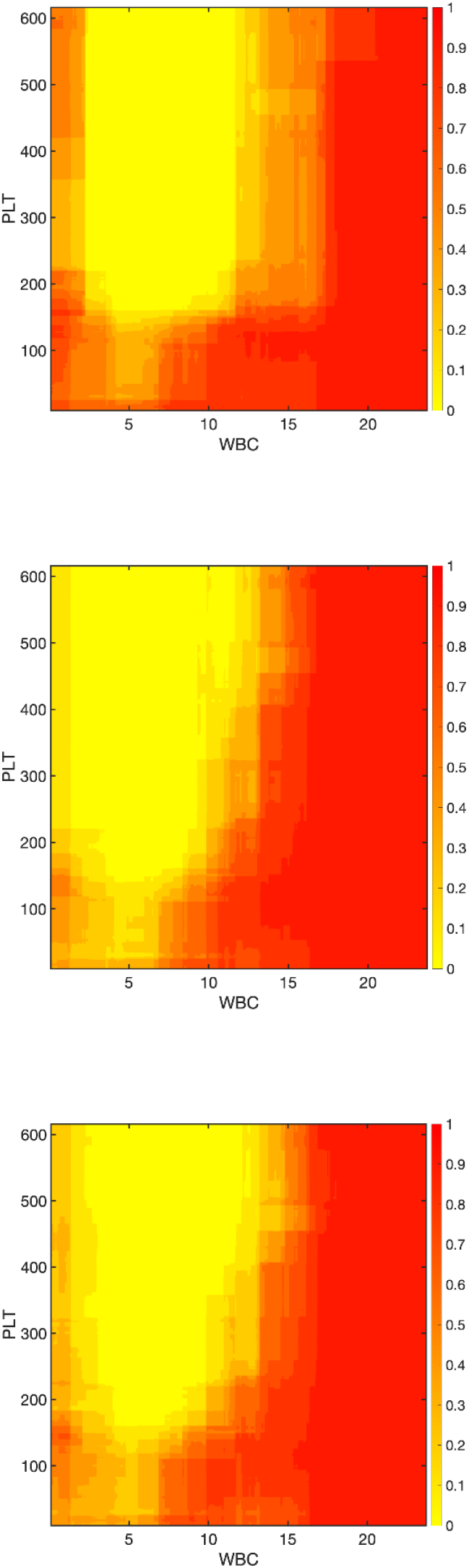
Heatmaps of model behaviour with all patients (1-3).

**Figure S10:**
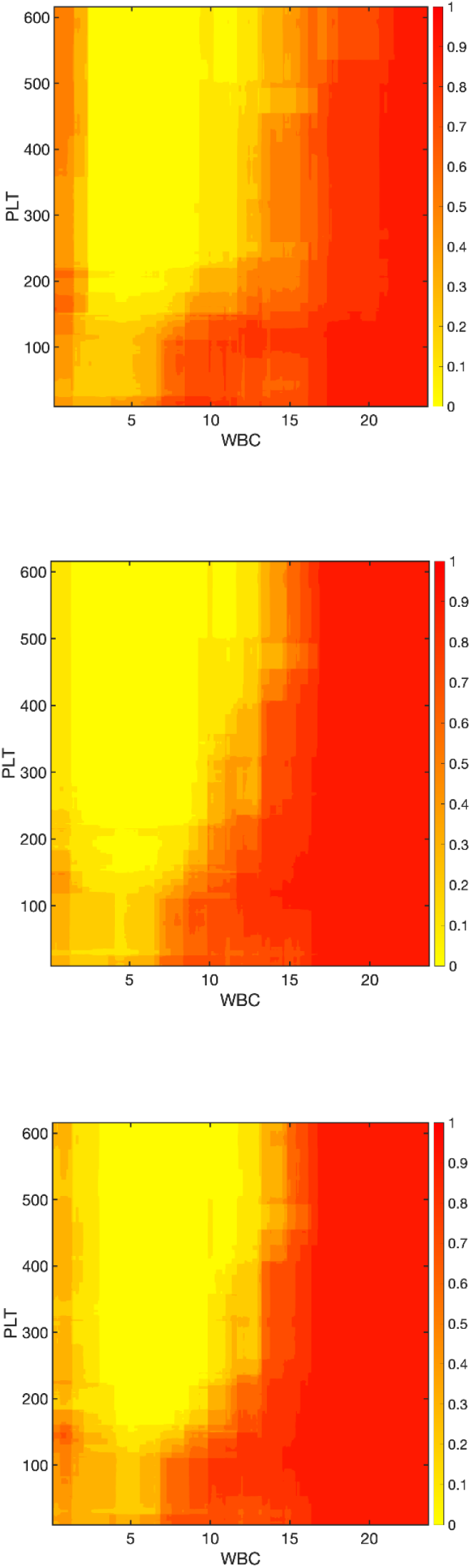
Heatmaps of model behaviour with sepsis patients (4-6).

## Notes

### Competing Interest Statement

The authors have declared no competing interest.

### Author Declarations

1. The Ethics Committee at the Leipzig University Faculty of Medicine approved the study (reference number: 214/18-ek). 2. The Ethics Committee at the University Medicine Greifswald approved the study (reference number: BB133/10).

